# Assessing Durability of Vaccine Effect Following Blinded Crossover in COVID-19 Vaccine Efficacy Trials

**DOI:** 10.1101/2020.12.14.20248137

**Authors:** Dean Follmann, Jonathan Fintzi, Michael P. Fay, Holly E. Janes, Lindsey Baden, Hana El Sahly, Thomas R. Fleming, Devan V. Mehrotra, Lindsay N. Carpp, Michal Juraska, David Benkeser, Deborah Donnell, Youyi Fong, Shu Han, Ian Hirsch, Ying Huang, Yunda Huang, Ollivier Hyrien, Alex Luedtke, Marco Carone, Martha Nason, An Vandebosch, Honghong Zhou, Iksung Cho, Erin Gabriel, James G. Kublin, Myron S. Cohen, Lawrence Corey, Peter B. Gilbert, Kathleen M. Neuzil

**Author notes:** Address reprint requests to Dr. Follmann at Biostatistics Research Branch, National Institute of Allergy and Infectious Diseases, National Institutes of Health, 5601 Fishers Lane, MSC 9820, Rockville, MD 20892-9820.

## Abstract

**Background:** Several candidate vaccines to prevent COVID-19 disease have entered large-scale phase 3 placebo-controlled randomized clinical trials and some have demonstrated substantial short-term efficacy. Efficacious vaccines should, at some point, be offered to placebo participants, which will occur before long-term efficacy and safety are known.

**Methods:** Following vaccination of the placebo group, we show that placebo-controlled vaccine efficacy can be derived by assuming the benefit of vaccination over time has the same profile for the original vaccine recipients and the placebo crossovers. This reconstruction allows estimation of both vaccine durability and potential vaccine-associated enhanced disease.

**Results:** Post-crossover estimates of vaccine efficacy can provide insights about durability, identify waning efficacy, and identify late enhancement of disease, but are less reliable estimates than those obtained by a standard trial where the placebo cohort is maintained. As vaccine efficacy estimates for post-crossover periods depend on prior vaccine efficacy estimates, longer pre-crossover periods with higher case counts provide better estimates of late vaccine efficacy. Further, open-label crossover may lead to riskier behavior in the immediate crossover period for the unblinded vaccine arm, confounding vaccine efficacy estimates for all post-crossover periods.

**Conclusions:** We advocate blinded crossover and continued follow-up of trial participants to best assess vaccine durability and potential delayed enhancement of disease. This approach allows placebo recipients timely access to the vaccine when it would no longer be proper to maintain participants on placebo, yet still allows important insights about immunological and clinical effectiveness over time.

---

Safe and durably effective SARS-CoV-2 vaccines hold the potential to dramatically alter the COVID-19 pandemic.^1,2^ Multiple vaccine candidates are currently in phase 3 placebo-controlled clinical testing with key criteria for success including vaccine efficacy and safety. Early results from at least three trials^3-5^ suggest high efficacy that far exceeds the FDA guidance threshold of 50% for symptomatic disease and severe disease. Yet critical questions remain, including the effects in subgroups such as the elderly and minorities and the assessment of longer-term efficacy and safety, given theoretical concerns for both vaccine associated enhanced disease (VAED; a potential concern in subgroups such as the elderly^6-8^) and waning protection.^6,9,10^ The latter concern must be considered in light of studies of seasonal coronaviruses^11-13^ and natural infection by SARS-CoV-2,^14-17^ which suggest immunity may wane within 6 months to 2 years in some people. Understanding whether durability of vaccine efficacy might be improved by revaccination is another critical question.^10,12,13^

Current COVID-19 phase 3 vaccine trials are large and designed to accrue at least 150 cases of clinical disease in a short period of time.^18-21^ For example, the mRNA coronavirus vaccine efficacy trials reached these endpoints within four months after trial initiation, reflecting the magnitude of the COVID-19 pandemic. While long-term safety and durability of efficacy are best evaluated by continued blinded follow-up of the original arms,^22^ at some point placebo volunteers must be offered the vaccine. The timing of this offer is complex with individual risk being weighed against the social value of the additional information, society’s perception of fairness, and the availability of the vaccine outside the trial.^23^ Ultimately, the participant’s view of equity will impact retention and determine how long blinded placebo control continues. Intuitively, placebo vaccination seems to preclude assessment of long-term comparative efficacy and VAED, as all are vaccinated. However, following crossover, the randomized trial remains, though now as a comparison of immediate (original vaccine arm) versus deferred (original placebo arm) vaccination. This allows assessment of waning vaccine effectiveness or associated enhanced disease. If, following crossover, the original vaccine arm accrues more cases than the original placebo arm, efficacy will be shown to wane. Furthermore, placebo-controlled vaccine efficacy can still be estimated after crossover, provided the trial continues with high levels of compliance and follow-up. To ensure the highest quality trial, blinded follow-up post crossover is ideal, wherein placebo recipients receive vaccine and vaccine recipients receive placebo.

Besides assessment of durability, continued follow-up allows for measurement of post-vaccination immune response in the newly vaccinated placebo recipients which can substantially increase the statistical power to assess immune correlates of risk. Continued follow-up readily allows a pivot to a randomized trial of a ‘booster’ dose of the vaccine to assess whether waning efficacy can be reversed.

## SETUP

A schematic for a blinded crossover trial is illustrated in Figure 1. Following initial equal randomization to vaccine or placebo, volunteers are followed for case accrual and at some point an efficacy signal is reached. After regulatory approval, blinded crossover occurs so that all willing volunteers have received the efficacious vaccine. At the point of crossover, the trial has changed into a blinded randomized trial of immediate (original vaccine) vs deferred (original placebo) vaccination. Following crossover there remain two distinct interventions that can be contrasted. Intuitively, if the placebo case accrual rate for the original vaccine arm is higher than the recently vaccinated, efficacy must be waning. Additionally, post-crossover vaccine efficacy can be estimated by assuming the newly vaccinated in period 1 or period 2 receive the same benefit. Figure 2 illustrates the concept for a vaccine with 80 % efficacy in period 1. We deduce the case count for an inferred period 2 placebo group by requiring that the inferred count align with the vaccine efficacy just observed in period 1 for the original vaccine recipients. We then use this inferred case count to deduce a period 2 vaccine efficacy. We note that the approach generalizes beyond two equal periods, but the key aspects are easiest to develop in this simpler setting.

**Figure 1.**
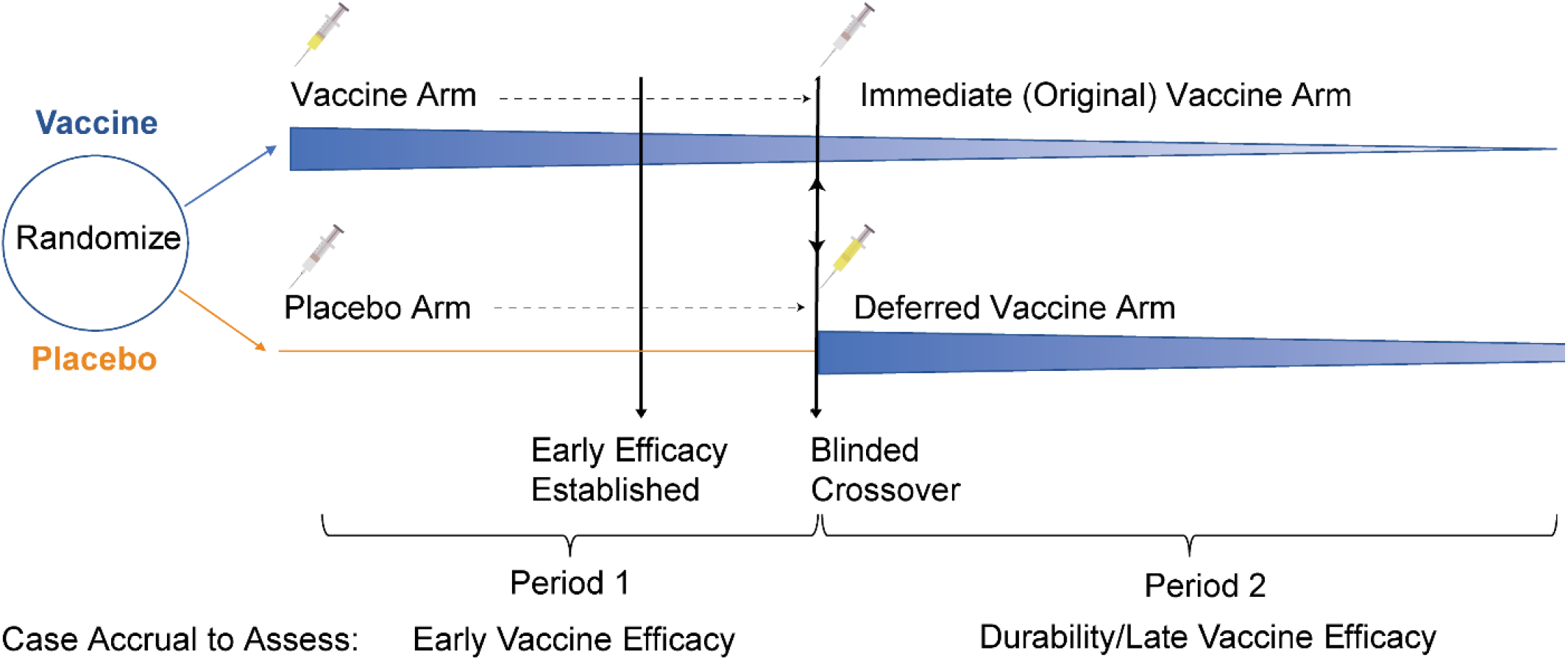
Schematic of a standard trial of vaccine vs placebo that pivots to a blinded crossover trial of immediate vs deferred vaccination. The tapering and fading blue wedge following vaccination evokes a potential waning of efficacy. At some point following a positive primary efficacy signal, placebo volunteers receive the vaccine and vaccine volunteers receive placebo. A balanced case split between arms in period 2 supports maintenance of the period 1 vaccine efficacy. A key assumption is that vaccine efficacy for the newly vaccinated is the same whether at the start of period 1 or at the start of period 2.

**Figure 2.**
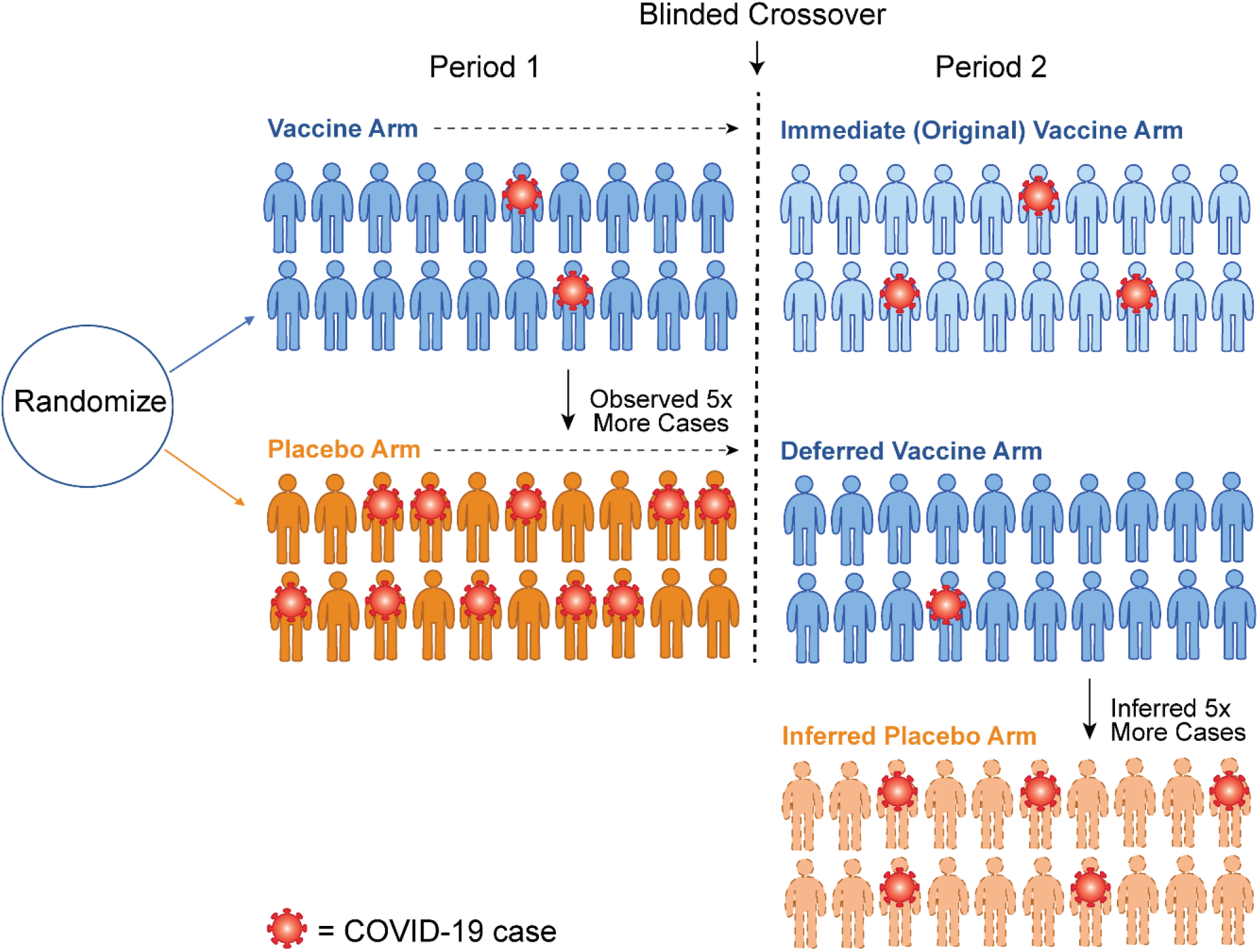
Schematic of how crossover allows imputation of the case counts for an inferred placebo group. Following crossover, we assume the vaccine efficacy in Period 2 for the newly vaccinated (Deferred Vaccine Arm) is the same 80% that was observed in the newly vaccinated (Immediate Vaccine Arm) in Period 1. This logic implies that a counterfactual placebo group of 20 volunteers would have about 5 cases. Thus the vaccine efficacy for the original vaccine arm in Period 2 has waned to 100% (1 – 3/5) = 60%.

Table 1 provides more detail. During period 1, vaccine efficacy is estimated as VE_1_ = 100% (1-25/125) = 80%, the reduction in observed cases as a result of the vaccine. Following crossover, we evaluate two scenarios: in scenario 1, the number of cases is similar in period 2 with a relative risk (original vaccine/original placebo) in period 2 of RR_2_= 41/39, which suggests little vaccine efficacy has been lost. Using the period 1 vaccine efficacy, we infer the number of placebo cases, that could have been seen, had there been a standard (non-crossover) trial, as 39 x (125/25) = 195. We then calculate vaccine efficacy in period 2 as VE_2_ = 100% 100%(1-41/195) =100%(1-[25/125] x [41/ 39]) = 100% (1-RR _1_ x RR _2_) = 79%. Note we obtain the same estimate of 79% even if the period 2 case counts are halved (or tripled) and thus this procedure does not require a constant placebo case accrual rate. For scenario 2, there is clear evidence of loss of durability as RR_2_= 53/9=5.89. Indeed, the vaccine efficacy becomes negative suggesting potential harm in period 2. Such a scenario is extreme, but does show how crossover could be used to assess late VAED.

**Table 1:** Illustrative data to demonstrate how a placebo-controlled vaccine efficacy in Period 2 can be calculated by inferring a placebo group in Period 2 following crossover for two different scenarios. In scenario 1, there is no waning of effect, whereas in scenario 2 the efficacy has waned for those originally vaccinated. In both scenarios, the vaccine efficacy in Period 1 for the newly vaccinated of 80% is assumed to apply to the newly vaccinated in Period 2. Bolded numbers are post vaccination cases, italicized numbers are inferred placebo cases.

| Arm | Period 1 | Crossover | Period 2<br>Scenario 1 | Period 2<br>Scenario 2 |
| --- | --- | --- | --- | --- |
| Arm | # cases |  | # cases | # cases |
| Original Vaccine<br>*(Immediate Vaccine)* | **25** |  | **41** | **53** |
| Original Placebo<br>*(Deferred Vaccine)* | 125 |  | **39** | **9** |
| Inferred Placebo | n/a | | $195=39 \times 125/25$ | $45=9 \times 125/25$ |
| Period Specific Vaccine<br>Efficacy<br>(95% Confidence Interval) | $80\%=1-25/125$<br>(69%, 88%) | | $79\%=1-41/195$<br>(60%, 89%) | $-18\%=1-53/45$<br>(-209%, 51%) |

## ASSUMPTIONS

A standard non-crossover trial requires 1) adequate retention following the original randomization such that the two randomized cohorts remain balanced at the start of period 2; 2) that the vaccine is similarly efficacious against circulating viruses in both periods; and 3) consistency in case assessment across periods, in order to obtain unbiased assessments about durability of efficacy. Importantly, the crossover design is valid without making assumptions about whether and how background incidence changes from period 1 to period 2. The only additional assumption required of the blinded crossover design is that the newly vaccinated obtain the same benefit of vaccination in period 1 as in period 2. This is implicitly assumed in all vaccine trials with rolling enrollment.

With an effective vaccine, the two groups might not be similar at the start of period 2, as the more vulnerable (e.g., riskier behavior or frailer health) placebo volunteers will acquire COVID-19 during period 1 at a greater rate than the matching vulnerable population of vaccinated volunteers, who were protected by vaccination.^24-27^ So period 2 can start with fewer vulnerable people in the original placebo arm compared to the original vaccine arm. Thus, the vaccine efficacy may appear to wane, but only because of a biased comparison. For large COVID-19 vaccine trials with relatively few cases, the fraction of participants excluded because of an event and resulting bias may not be a substantial concern. Additionally, true waning vaccine efficacy is supported if the kinetics of antibody waning in individuals track with individual risk over time, so long as the infecting pathogen does not undergo evolution that renders it less susceptible to the vaccine.^28^

## ESTIMATION AND COMPARATIVE PERFORMANCE

The modified Poisson method to estimate overall vaccine efficacy can be generalized to provide period-specific estimates of relative risk and vaccine efficacy.^29,30^ This person-year approach allows for differential follow-up within each period due to rolling enrollment or dropout. More than two periods with different durations can be assessed and with sophisticated methods, a period-free curve of placebo-controlled vaccine efficacy throughout follow-up can be derived.

Table 2 evaluates the performance of a standard non-crossover trial compared to a crossover trial in terms of power or the probability of detecting waning efficacy and the power of detecting harm in period 2, with calculations implemented in the R package plaXdesign.^31^ We also report sample size ratios, where for example, a sample size ratio of 2 means a crossover trial would require twice as many volunteers to achieve the power of a given standard trial. The first four rows correspond to a scenario with high initial efficacy that accrues about 200 COVID-19 cases in placebo recipients during period 1 and accrues 200 or 100 COVID-19 cases in placebo recipients during period 2 in a standard trial. Under crossover these period 2 placebo cases are inferred. We have good power to detect waning efficacy when VE changes from 90% to 75% with 200 cases in period 2 under either the crossover (0.93 power) or standard design (0.90 power). Power is lessened with 100 cases in period 2 with crossover having 0.69 power and standard having 0.81 power.

**Table 2:**
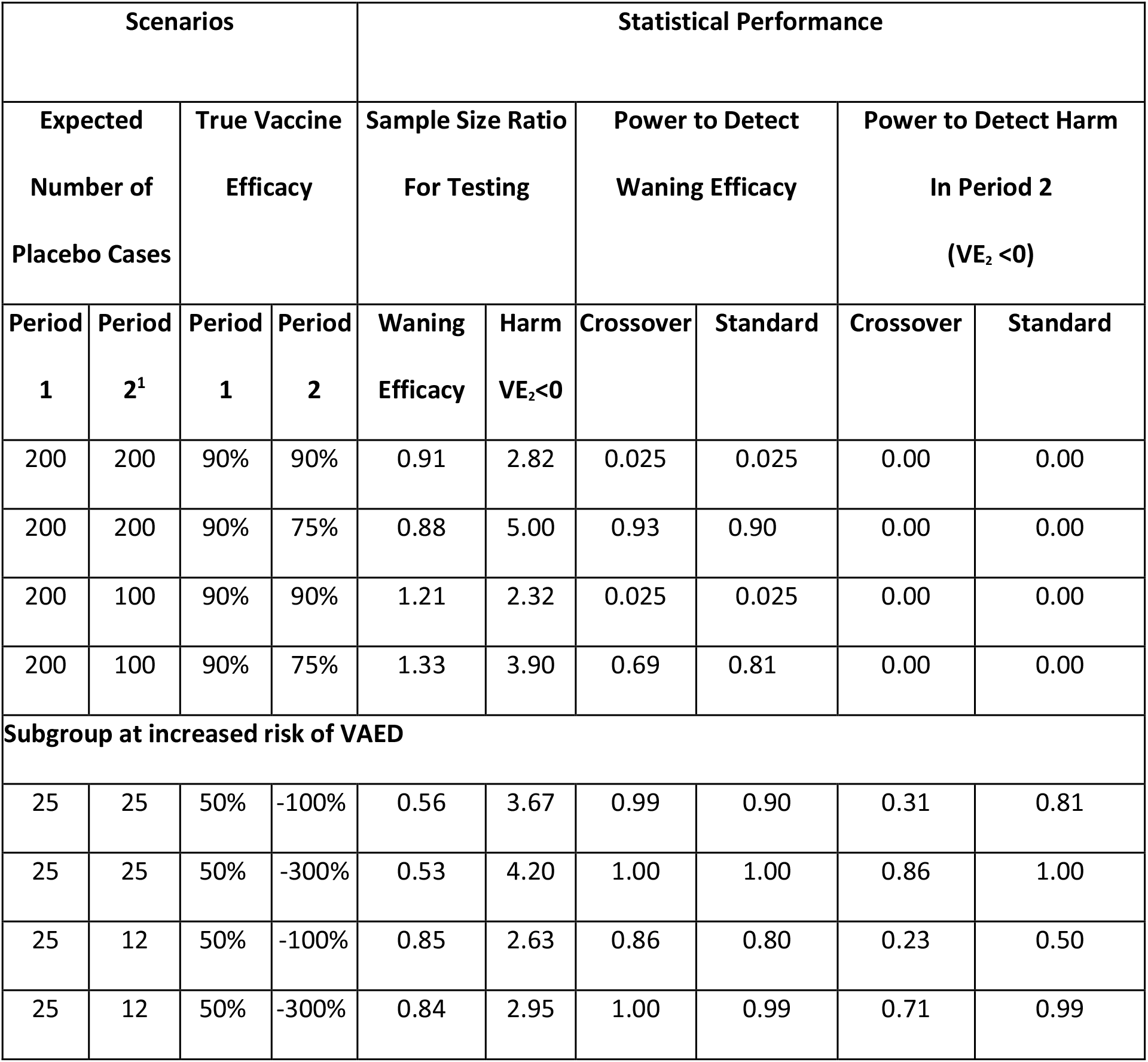
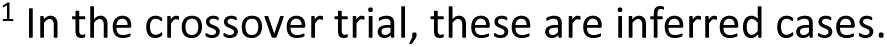
Statistical performance of the crossover design compared to the standard non-crossover design. The first four rows denote scenarios with high efficacy where waning efficacy is of major interest. The second four rows correspond to a subgroup where efficacy is more modest and vaccine harm or VAED is of greater concern. A sample size ratio of 2 means a crossover trial would need to have twice the sample size to achieve the same power as a standard trial. VE_2_ is the vaccine efficacy in period 2. A README is available at the plaXdesign page on Github^31^ with instructions on how to reproduce this table.

| Scenarios |  |  |  | Statistical Performance |  |  |  |  |  |
| --- | --- | --- | --- | --- | --- | --- | --- | --- | --- |
| Expected Number of Placebo Cases | | True Vaccine Efficacy | | Sample Size Ratio For Testing | | Power to Detect Waning Efficacy | | Power to Detect Harm In Period 2 ($VE_2 < 0$) | |
| Period 1 | Period 2<sup>1</sup> | Period 1 | Period 2 | Waning Efficacy | Harm $VE_2 < 0$ | Crossover | Standard | Crossover | Standard |
| 200 | 200 | 90% | 90% | 0.91 | 2.82 | 0.025 | 0.025 | 0.00 | 0.00 |
| 200 | 200 | 90% | 75% | 0.88 | 5.00 | 0.93 | 0.90 | 0.00 | 0.00 |
| 200 | 100 | 90% | 90% | 1.21 | 2.32 | 0.025 | 0.025 | 0.00 | 0.00 |
| 200 | 100 | 90% | 75% | 1.33 | 3.90 | 0.69 | 0.81 | 0.00 | 0.00 |
| **Subgroup at increased risk of VAED** |  |  |  |  |  |  |  |  |  |
| 25 | 25 | 50% | -100% | 0.56 | 3.67 | 0.99 | 0.90 | 0.31 | 0.81 |
| 25 | 25 | 50% | -300% | 0.53 | 4.20 | 1.00 | 1.00 | 0.86 | 1.00 |
| 25 | 12 | 50% | -100% | 0.85 | 2.63 | 0.86 | 0.80 | 0.23 | 0.50 |
| 25 | 12 | 50% | -300% | 0.84 | 2.95 | 1.00 | 0.99 | 0.71 | 0.99 |

<sup>1</sup> In the crossover trial, these are inferred cases.

The bottom four rows focus on a scenario where a subgroup may be at risk of VAED. The subgroup accrues about 25 placebo cases in period 1 thus about 1/8 of the total of 200 placebo cases. As before, we anticipate around 25 or 12 placebo cases in period 2 under the standard trial. We consider subgroup vaccine efficacies in Period 2 of –100% and –300% or a doubling and quadrupling of the case rate on vaccine compared to a placebo group, respectively. These estimates are meant to roughly parallel the vaccine-enhanced risk of hospitalized dengue disease and of severe dengue disease seen with the CYD-TDV dengue vaccine in baseline-seronegative participants.^32^ Power to detect waning efficacy is greater than 0.80 for all scenarios. The power to detect harm is substantially worse under the crossover design compared to the standard trial with poor power to detect harm with a period 2 VE of –100%, but powers of 0.71 and 0.85 to detect harm with a VE of –300% with 12 and 25 expected cases in period 2, respectively.

Figure 3 demonstrates the behavior of the method for more than two periods. The vaccine efficacy estimates for post-crossover periods depend on all prior VE estimates so if any period has an unreliable estimate of VE, so will subsequent periods. Evaluation of waning efficacy, within any post-crossover period is a simple contrast of two randomized arms and, with sufficient cases and blinded follow-up, can be rigorously evaluated, even after a ‘weak link’ period. Figure 3 suggests that longer periods (longer links) with higher case counts (wider links) will provide better estimates of late vaccine efficacy. This underscores the statistical benefit of maintaining the original placebo-controlled design for as long as possible to maximize the first period.

**Figure 3.**
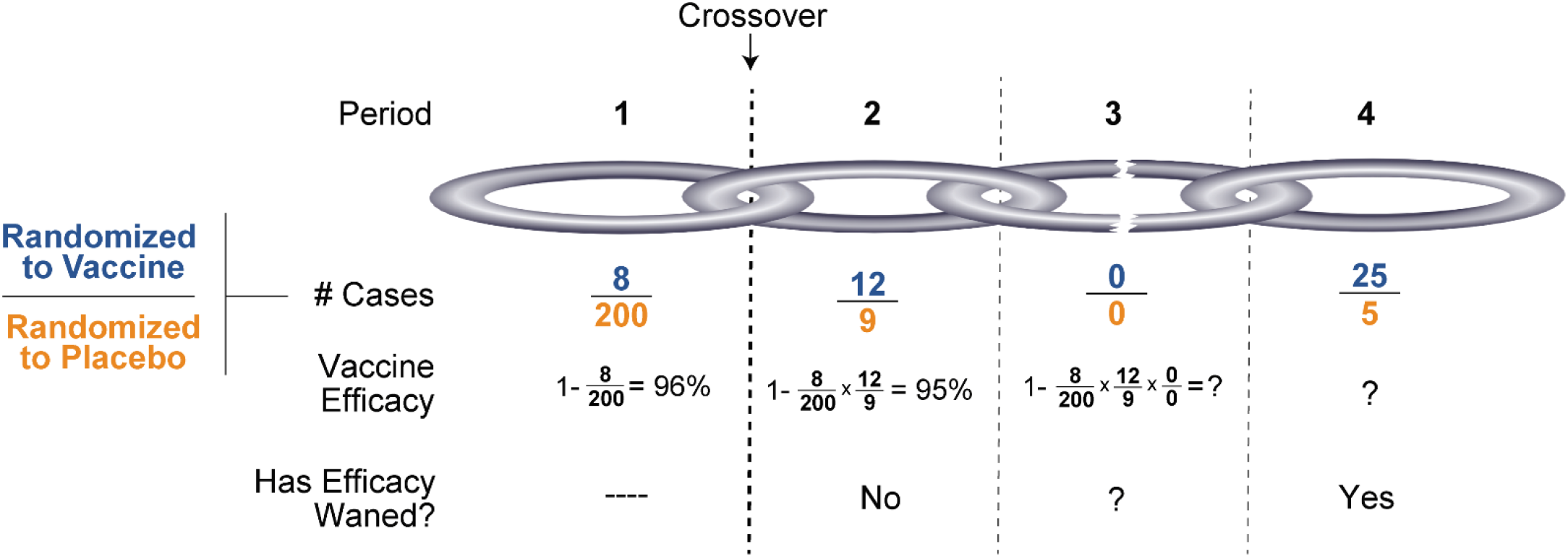
Following crossover, the period-specific vaccine efficacy estimates depend on all previous periods. A period with few cases makes subsequent efficacy estimates unstable but paradoxically does not preclude evaluation of subsequent waning efficacy. A bigger and stronger chain is achieved with a long first (and thus subsequent) period and more cases within each period.

## COLLATERAL BENEFITS OF PLACEBO CROSSOVER

Correlates of risk and correlates of protection analyses involve describing associations between the measured immune response shortly after the last dose of vaccine and subsequent risk of COVID-19 disease.^33-35^ A proven or “reasonably likely” immune correlate of protection can allow for immunobridging to populations not included in the trials either through traditional or provisional approval, for example through the FDA’s Accelerated Approval mechanism.^36,37^ This makes the establishment of correlates of risk and protection a key goal of vaccines trials. Following a strong positive efficacy result, an immune correlates analysis may be underpowered due to relatively few breakthrough cases in the vaccine recipients. However, placebo crossover could effectively double the sample size for assessing immune correlates of risk and protection, a notable benefit of crossover.

If vaccine efficacy wanes substantially over the course of follow-up, a natural question will be whether revaccination can reverse the loss. Continued follow-up of the volunteers provides a ready-to-go vehicle to experimentally evaluate revaccination by randomizing those who received vaccine to another course of vaccine or placebo. While a boost trial can be quickly conducted in a standard trial that maintains follow-up, placebo crossover allows for a doubling of those who are available for the boost trial relative to a trial that maintains a placebo control. A boost trial following crossover could proceed in two stages---the original vaccine arm would be randomized first and the crossover placebos randomized later, if needed (Figure A1 in the Appendix). If the vaccine efficacy wanes from 80% to 40% and it is hoped boosting will recover the VE to 80%, about 42 cases are required to achieve 90% power, substantially fewer than the roughly 150 cases required for the original trial.

## IMPLEMENTATION

Once the vaccine is available, principles of ethics (including an evaluation of the social benefit and individual risk profile of the trial), transparency, and fairness should govern the timing and implementation of the planned crossover.^23^ Ideally a blinded crossover phase would be an integral part of the original design and part of the initial consent. Otherwise, a protocol amendment would be introduced and reconsent obtained.

Performing the crossover blinded offers substantial scientific advantages. Just as the pre-crossover period was blinded to address initial vaccine efficacy, so should the post-crossover period be blinded to address durability. Both questions are critical and deserve the same rigor. With an unblinded crossover, there is real concern that immediately after crossover the newly unblinded original vaccine recipients will forgo masks and social distancing while the newly unblinded placebo volunteers will not. Another concern is that mild subjective symptoms might be differentially dismissed/elevated in the unblinded arms. This is particularly important since efficacy is entirely predicated on volitional presentation with signs or symptoms of COVID-19. Such differential behavior would substantially confound a between-arm case split consistent with waning efficacy in period 2. Furthermore, such a ‘biased link’, as illustrated in Figure 3, would confound interpretation of all later periods. With minimal assessment of safety and reactogenicity in the newly vaccinated, operationally maintaining the blind could essentially only require the addition of dummy shot(s) in the original vaccine recipients with some additional blood draw(s). Nonetheless, if some volunteers choose to become unblinded, continued follow-up of those participants should be pursued to maximize information. Simple questions to assess planned and actual risk behavior, along with comorbidities, could help adjust for the biases caused by unblinding.

An important goal for COVID-19 vaccine trials is to assess the effect of vaccine on asymptomatic infection as assessed by periodic serologies. To maintain this goal, serologies should be collected in all volunteers at the time of crossover with both arms having the same schedules post crossover to ensure even-handed evaluation of this endpoint. While more complex, the methods of this paper can be generalized to study vaccine efficacy against infrequently assessed endpoints such as seroconversion.

Different approaches to crossing over could be implemented, with a key requirement being that both arms are treated the same. The simplest would be to plan for all participants to crossover. If the vaccine is initially recommended for a subgroup, e.g. higher risk individuals, crossover might initially target the higher risk subgroup with the blinded placebo-controlled trial continuing for the lower risk subgroup. Crossover could later occur for the lower risk subgroup if an efficacy signal is achieved. Operationally, crossover could follow the order of the original enrollment but occur at an accelerated pace compared to enrollment, so that different cohorts would crossover on different days relative to the initial vaccination. Another possibility is to randomize the time of crossover. To reduce volunteer fatigue, late follow-up might be lessened provided critical endpoints are still assessed as in the pre-crossover period.

## CONCLUSIONS

The high efficacies reported in primary^3^ and interim^4,5^ analyses of multiple vaccine candidates, while universally welcomed, add complexity and uncertainty to the environment surrounding access to the vaccine for trial participants randomized to placebo. Continued blinded follow-up in the original arms is optimal to assess vaccine efficacy over time and is endorsed by the FDA in their guidance pertaining to COVID-19 vaccine development. Early efficacy provides incomplete information about the totality of the risks and benefits of the vaccines. But at some point, consensus will emerge that the placebo volunteers should be offered vaccine. This paper argues that valuable information regarding durability and VAED can be obtained even after the placebo volunteers receive the vaccine and that studies should maintain rigorous blinded follow-up post crossover to recover this information. Additionally, continued follow-up allows for a doubling of the volunteers who can contribute to an immune correlates analysis and allows for a quick pivot to a trial of boosting should the vaccine demonstrate waning efficacy.

### Grant Support

This work was supported by grants from the National Institute of Allergy and Infectious Diseases of the National Institutes of Health, through grant numbers UM1AI068635, R37AI054165, UM1AI068617, UM1Al148684, and R37-AI29168. The content is solely the responsibility of the authors and does not necessarily represent the official views of the National Institutes of Health.

## Data Availability

All data referred to in the manuscript are included within the manuscript and its appendix.

