## Appendix for "Assessing Durability of Vaccine Effect Following Blinded Crossover in COVID-19 Vaccine Efficacy Trials"

**Recovery of VE in period 2 under crossover**

We begin by demonstrating how the placebo-controlled vaccine efficacy in period 2, is obtainable following placebo crossover. Denote the expected number of cases in arm Z= 1 (original vaccine), 0 (original placebo) in period k=1,2 by $\theta_{ZK}$ and $\omega_{ZK}$, for the standard and crossover trial respectively. For the standard trial, we define the relative risk of original vaccine to original placebo in period k as $RRS_K = \theta_{1K}/\theta_{0K}$ and define the vaccine efficacy as $VE_K = 1 - RRS_K$. For the crossover trial we define the relative risk in period k as $RRX_K = \omega_{1K}/\omega_{0K}$. Since period 1 and the original vaccine arm in period 2 are identical under both designs we have $\theta_{Z1} = \omega_{Z1}$, $\theta_{12} = \omega_{12}$ and $RRX_1 = RRS_1$. We make the assumption that $VE_1$ applies to both the newly vaccinated during period 1 and the newly vaccinated during period 2. Since $VE_1 = 1 - RRX_1$, $RRX_1$ also applies to the newly vaccinated in period 2. In the standard trial, $VE_2 = 1 - RRS_2$. To recover $VE_2$ in the crossover trial note that the true crossover counterfactual placebo rate in period 2 is $(1/RRX_1)\,\omega_{02}$. Now

$$VE_2 = 1 - \theta_{12}/\theta_{02} = 1 - \omega_{12}/\{(1/RRX_1)\,\omega_{02}\} = 1 - RRX_1\omega_{12}/\omega_{02} = 1 - RRX_1 RRX_2$$

Thus, to recover the VE in period 2 of a standard trial when confronted with a placebo crossover trial we simply take one minus the product of the relative risks for the two periods of the crossover. Table A1 shows the connection between the parameters of the Standard and Crossover Trials.

**Table A1: Expected cases counts for the different arms and periods under the standard and crossover trials.**

| | Standard Trial | | Crossover Trial | |
|---|---|---|---|---|
| | **Period 1** | **Period 2** | **Period 1** | **Period 2** |
| Original Vaccine | $\theta_{11} = \theta_1(1\text{-}VE_1)$ | $\theta_{12} = \theta_2(1\text{-}VE_2)$ | $\omega_{11} = \theta_1(1\text{-}VE_1)$ | $\omega_{12} = \theta_2(1\text{-}VE_2)$ |
| Original Placebo | $\theta_{01} = \theta_1$ | $\theta_{02} = \theta_2$ | $\omega_{01} = \theta_1$ | $\omega_{02} = \theta_2(1\text{-}VE_1)$ |

**Efficiency of estimation and power for testing harm and waning VE under the two designs**

Vaccine durability means that $VE_1 = VE_2$. Assessin vaccine durability is equivalent to assessing whether $(1- VE_2)/ (1 - VE_1) = RRS_2/RRS_1 = 1$, a ratio of two ratios. Under crossover, assessing durability is equivalent to assessing whether $(1- VE_2)/ (1 - VE_1) = (RRX_1 RRX_2)/ RRX_1 = RRX_2 = 1$, a single ratio.

We next derive simple expressions for the variance of the vaccine efficacy estimates in each period. We assume that disease acquisition is rare so that $Y_{ZK}$, the number of cases in original arm Z =0, 1 in period K=1,2 is Poisson with parameter $\theta_{ZK}$ under the standard design. We analogously assume $X_{ZK}$, the number of cases in original arm Z =0, 1 in period K=1,2 is Poisson with parameter $\omega_{ZK}$ under the crossover design. Note $Y_{ZK} = X_{ZK}$ for (Z,K)=(1,1), (0,1), and (1,2), but $Y_{02}$ has a different distribution from $X_{02}$. To estimate the vaccine efficacies or relative risks, we simply take ratios of case counts of the different periods. For example, $\widehat{VES}_2 = 1 - Y_{12}/Y_{02}$ and $\widehat{VEX}_2 = (1 - [X_{11}/X_{01}] \times [X_{12}/X_{02}])$ under the standard and crossover trials, respectively. Using the delta method, one can show that the ratio of variances for the standard versus crossover estimates is approximately

$$\text{RVE2} = \frac{\text{var}\{\log(1-\widehat{VES}_2)\}}{\text{var}\{\log(1-\widehat{VEX}_2)\}} =$$

$$\left(\frac{1}{\theta_2} + \frac{1}{\theta_2(1-VE_2)}\right) / \left(\frac{1}{\theta_1} + \frac{1}{\theta_1(1-VE_1)} + \frac{1}{\theta_2(1-VE_1)} + \frac{1}{\theta_2(1-VE_2)}\right).$$

Note that as $\theta_1$ becomes large, RVE2 approaches 1 (if $VE_1$=0 and $\theta_2$ remains constant). This underscores the advantage of having a large number of events in period 1 in order to maximize the efficiency of the crossover design relative to the standard design to estimate vaccine efficacy in period 2 and to assess harm.

The ratio of variances for the estimate of the period 1 to period 2 relative risk is given by

$$\text{RVE12} = \frac{\text{var}[\log\{(1-\widehat{VES}_1)/(1-\widehat{VES}_2)\}]}{\text{var}[\log\{(1-\widehat{VEX}_1)/(1-\widehat{VEX}_2)\}]} =$$

$$\left(\frac{1}{\theta_1} + \frac{1}{\theta_1(1-VE_1)} + \frac{1}{\theta_2} + \frac{1}{\theta_2(1-VE_2)}\right) / \left(\frac{1}{\theta_2(1-VE_1)} + \frac{1}{\theta_2(1-VE_2)}\right).$$

To interpret the ratios RVE2 and RVE12, we note that in terms of estimation efficiency, a standard trial with N subjects that achieved a given power would require N/RVE subjects to achieve the same power under a crossover trial. So these ratios provide a simple way to evaluate the relative statistical performance of the two designs when estimating either $VE_2$ or $VE_1/VE_2$ .

For power calculations, note that for an alpha=.025 one-sided test that $\mu \leq 0$, the power of a asymptotically normal test statistic with mean $\mu$ and variance 1 is approximately given by

$$\Phi(\mu - 1.96),$$

while the analogous test that $\mu \geq 0$ is given by $\Phi(-\mu - 1.96)$, where $\Phi$ is the standard normal cumulative distribution function.

Table A2 provides the $\mu$ for tests of waning efficacy and harm under the standard and placcebo crossover designs. The actual test statistics would replace the parameters with estimated parameters, i.e. case counts.

**Table A2: The mean of asymptotically normal tests of waning efficacy and harm under standard and crossover designs. The parameters $\theta_K$ and $VE_K$ denote the expected number of placebo cases in period K and the vaccine efficacy in period K, respectively. The tests have unit variance.**

| Test | $\mu$ Standard Trial | $\mu$ Crossover Trial |
|---|---|---|
| Waning Efficacy H0: $VE_1 = VE_2$ | $\{\log(1-VE_1) - \log(1-VE_2)\}/\sqrt{\frac{1}{\theta_1}+\frac{1}{\theta_1(1-VE_1)}+\frac{1}{\theta_2}+\frac{1}{\theta_2(1-VE_2)}}$ | $\{\log(1-VE_1) - \log(1-VE_2)\}/\sqrt{\frac{1}{\theta_2(1-VE_1)}+\frac{1}{\theta_2(1-VE_2)}}$ |
| Harm H0: $VE_2 = 0$ | $\log(1-VE_2)/\sqrt{\frac{1}{\theta_2}+\frac{1}{\theta_2(1-VE_2)}}$ | $\log(1-VE2)/\sqrt{\frac{1}{\theta_1}+\frac{1}{\theta_1(1-VE_1)}+\frac{1}{\theta_2(1-VE_1)}+\frac{1}{\theta_2(1-VE_2)}}$ |

**Parameter Estimation**

We finish by noting that estimation of the relative risks and vaccine efficacies can be accomplished by using Generalized Estimating Equations aka modified Poisson regression [25,30]. For each subject we define $Y_1$, $T_1$, Z which are the indicator of a case in period 1, the follow-up time in period 1 and the original vaccine indicator, respectively. $T_1$ equals the case time for a case in period 1, the time of dropout for dropouts in period 1, and the length of period 1 for those who are period 1 non-cases and followed past period 1. For subjects with follow-up in period 2 (i.e. period 1 non-cases with follow-up longer than period 1), we define $Y_2$, $T_2$ analogously. We can then fit the working Poisson model for the crossover trial for which

$$E(Y_K) = \exp\{\beta_0 + \beta_1 Z + \beta_2 I(K=2) + \beta_3 Z\, I(K=2) + \log(T_K)\}$$

The parameters can be estimated using Generalized Estimating Equations software with a sandwich variance estimator but treating the data across the two periods as independent. To illustrate use of the model we provide two examples. To obtain the estimate of period 2 vaccine efficacy under the placebo crossover trial we form

$$\widehat{VEX}_2 = 1 - \exp(\widehat{\beta_1}) \exp(\widehat{\beta_1} + \widehat{\beta_3}).$$

Under the standard trial the parameters associated with period 2 are different and we write

$$E(Y_K) \quad = \exp\{\alpha_0 + \alpha_1 Z + \alpha_2 I(K = 2) + \alpha_3 Z I(K = 2) + \log(T_K)\}$$

$$\widehat{VES}_2 = 1 - \exp(\widehat{\alpha_1} + \widehat{\alpha_3})$$

Baseline covariates that reflect the risk of case acquisition can be incorporated into the model both to improve the precision of the estimates as well as to ameliorate any bias due to greater removal of the riskier volunteers from the placebo group.

Our development has focused on two periods with equal duration with a constant VE in each period. This approach can be extended to multiple periods and a parameterized vaccine efficacy curve specified.

A more elegant approach to modeling is given by Cox regression with time-dependent covariate which completely obviates the need to specify periods and can readily allow for smoothly varying vaccine efficacy in addition to the piecewise constant vaccine efficacy of this paper. Development of this method is the focus of another paper.

Many vaccine trials use per-protocol type analyses where cases are not counted until sometime after the final dose has been administered. Such analyses can be addressed by not counting cases or follow-up during this period. For example if cases are counted day 43 post first dose, this 'black-out' period would apply to both arms and both for the original dosing and the crossover dosing.

**Small Sample Calculation of VE in a Placebo Crossover Trial**

Assume $X_{ZK}$, the number of cases in original arm Z =0, 1 in period K=1,2 is Poisson with parameter $\omega_{ZK}$ under the crossover design. For any period, condition on the total number of cases, so that

$X_{11} | X_{11} + X_{01} = t_1 \sim Binomial(t_1, \pi_1)$ and $X_{12} | X_{12} + X_{02} = t_2 \sim Binomial(t_2, \pi_2)$

Then

$$VE_2 = 100\left\{1 - \left(\frac{\pi_1}{1-\pi_1}\right)\left(\frac{\pi_2}{1-\pi_2}\right)\right\}.$$

This presentation assumes equal follow-up between the arms. Adjustments for unequal follow-up are not presented but can be done (for analogous adjustments, see Dragalin, et al, 2002, Section 4.1).

Fay, et al (2015) developed a frequentist confidence interval method for combining parameters based on independent observations. We treat $X_{11} | X_{11} + X_{01} = t_1$ and $X_{12} | X_{12} + X_{02} = t_2$ as independent binomials, and get melded confidence intervals for $VE_2$,

$$\left\{q\left(\frac{\alpha}{2}, 100\left\{1 - \left(\frac{B_{U1}}{1-B_{U1}}\right)\left(\frac{B_{U2}}{1-B_{U2}}\right)\right\}\right), q\left(1 - \frac{\alpha}{2}, 100\left\{1 - \left(\frac{B_{L1}}{1-B_{L1}}\right)\left(\frac{B_{L2}}{1-B_{L2}}\right)\right\}\right)\right\},$$

where

$$B_{LK} \sim Beta(x_{1K}, x_{0K} + 1)$$

$$B_{UK} \sim Beta(x_{1K} + 1, x_{0K})$$

For K=1,2, and q(a,W) is the ath quantile of any random variable W. This can be easily calculated with Monte Carlo methods, where simulations sizes of $10^6$ can give quick and very precise confidence limit estimates. This method is designed to guarantee coverage. It is a slight generalization of the melding method, simpler than the generalization of Fay and Lumbard, 2020. Compare to the Bayesian method using Jeffreys prior for the binomial, Beta(0.5,0.5), the expression is the same except with

$B_{LK} \equiv B_{UK} \sim Beta(x_{1K} + 0.5, x_{0K} + 0.5)$.

**Additional Evaluation**

The following two tables provide a statistical evaluation of the different methods when the expected number of placebo cases for period 1, period 2 is 200,400 and 400, 200 respectively.

**Table A3: Statistical evaluation of the placebo crossover design compared to the standard design. Periods 1 and 2 have equal length and an expected placebo case rate of 200 in period 1 and 400 in Period 2.**

| True Vaccine Efficacy | | Sample Size Ratio | | Power to Detect Waning Efficacy | | Power to Detect Harm In Period 2 | |
|---|---|---|---|---|---|---|---|
| **Period 1** | **Period 2** | **Waning Efficacy** | **$VE_2$** | **Placebo Crossover** | **Standard** | **Placebo Crossover** | **Standard** |
| 80% | 80% | 0.56 | 3.67 | 0.025 | 0.025 | 0.00 | 0.00 |
| 80% | 60% | 0.48 | 5.57 | 1.00 | 0.94 | 0.00 | 0.00 |
| 80% | 40% | 0.45 | 7.00 | 1.00 | 1.00 | 0.00 | 0.00 |
| 80% | -20% | 0.42 | 9.73 | 1.00 | 1.00 | 0.14 | 0.77 |
| 80% | -40% | 0.42 | 10.33 | 1.00 | 1.00 | 0.36 | 1.00 |
| 50% | 50% | 0.44 | 3.33 | 0.025 | 0.025 | 0.00 | 0.00 |
| 50% | 30% | 0.41 | 3.88 | 0.95 | 0.64 | 0.00 | 0.00 |
| 50% | 10% | 0.38 | 4.32 | 1.00 | 0.98 | 0.00 | 0.00 |
| 50% | -20% | 0.36 | 4.82 | 1.00 | 1.00 | 0.23 | 0.77 |
| 50% | -40% | 0.35 | 5.08 | 1.00 | 1.00 | 0.63 | 1.00 |

**Table A4: Statistical evaluation of the placebo crossover design compared to the standard design. Periods 1 and 2 have equal length and an expected placebo case rate of 400 in period 1 and 200 in Period 2.**

| True Vaccine Efficacy | | Sample Size Ratio | | Power to Detect Waning Efficacy | | Power to Detect Harm In Period 2 | |
|---|---|---|---|---|---|---|---|
| **Period 1** | **Period 2** | **Waning Efficacy** | **$VE_2$** | **Placebo Crossover** | **Standard** | **Placebo Crossover** | **Standard** |
| 80% | 80% | 1.11 | 2.17 | 0.025 | 0.025 | 0.00 | 0.00 |
| 80% | 60% | 1.15 | 3.00 | 0.95 | 0.97 | 0.00 | 0.00 |
| 80% | 40% | 1.18 | 3.63 | 1.00 | 1.00 | 0.00 | 0.00 |
| 80% | -20% | 1.21 | 4.82 | 1.00 | 1.00 | 0.14 | 0.48 |
| 80% | -40% | 1.21 | 5.08 | 1.00 | 1.00 | 0.36 | 0.95 |
| 50% | 50% | 0.89 | 1.83 | 0.025 | 0.025 | 0.00 | 0.00 |
| 50% | 30% | 0.87 | 2.03 | 0.73 | 0.67 | 0.00 | 0.00 |
| 50% | 10% | 0.86 | 2.18 | 1.00 | 0.99 | 0.00 | 0.00 |
| 50% | -20% | 0.85 | 2.36 | 1.00 | 1.00 | 0.24 | 0.48 |
| 50% | -40% | 0.84 | 2.46 | 1.00 | 1.00 | 0.64 | 0.95 |

Software is available to reproduce the calculations presented in the tables of this paper. For example, Table 2 can be reproduced using R package plaXdesign as follows:

```
library(devtools)
install_github("mjuraska/plaXdesign")
library(plaXdesign)
plaXpower(theta1=c(rep(200,4), rep(25,4)),
        theta2=c(rep(200,2), rep(100,2), rep(25,2), rep(12,2)),
        ve1=c(rep(0.9, 4), rep(0.5, 4)),
        ve2=c(0.9, 0.75, 0.9, 0.75, -1, -3, -1, -3))
```

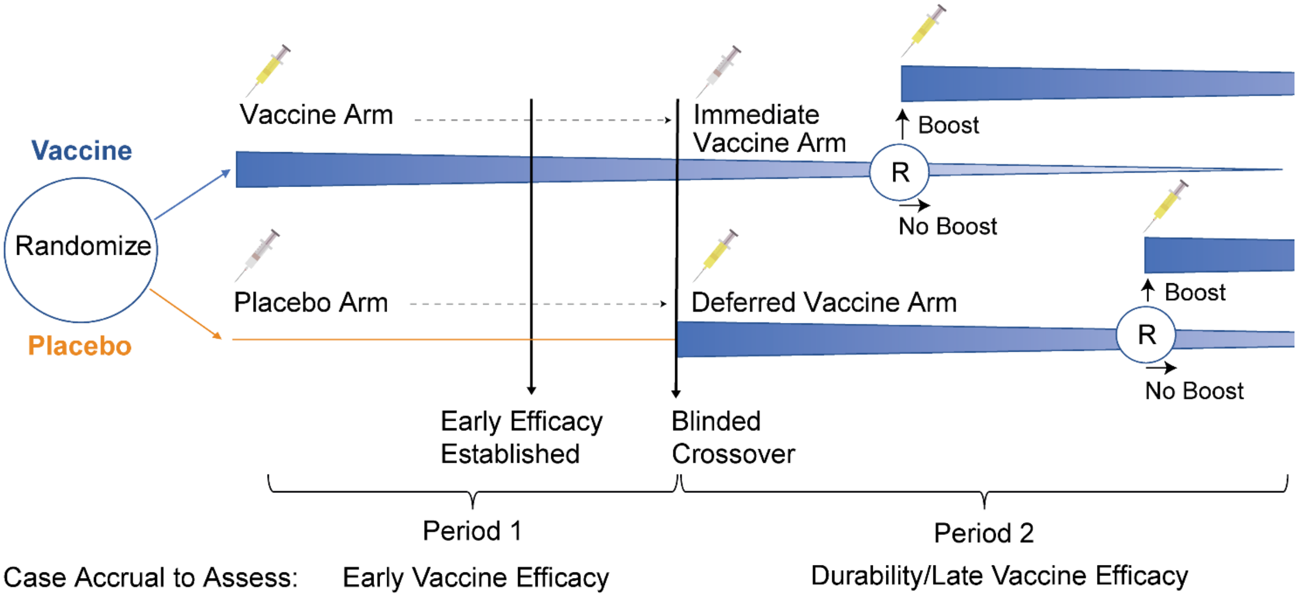

**Figure A1. Schematic of a vaccine vs placebo trial that pivots to a blinded crossover trial of immediate vs deferred vaccination with a boost.** The immediate (original) vaccine arm is randomized first and the crossover placebos (deferred vaccine arm) are randomized later, if needed.
